# Protocol for mixed-method study by LOng COvid Multidisciplinary consortium: Optimising Treatments and servIces acrOss the NHS (LOCOMOTION)

**DOI:** 10.1101/2022.04.09.22273655

**Authors:** Manoj Sivan, Trisha Greenhalgh, Julie L. Darbyshire, Ghazala Mir, Rory J. O’Connor, Helen Dawes, Darren C. Greenwood, Daryl B. O’Connor, Mike Horton, Stavros Petrou, Simon de Lusignan, Vasa Curcin, Erik Mayer, Alexander Casson, Ruairidh Milne, Clare Rayner, Nikki Smith, Amy Parkin, Nick Preston, Brendan Delaney, the LOCOMOTION CONSORTIUM

## Abstract

**Introduction:** Long COVID, a new condition whose origins and natural history are not yet fully established, currently affects 1.5 million people in the UK. Most do not have access to specialist long COVID services. We seek to optimise long COVID care both within and outside specialist clinics, including improving access, reducing inequalities, helping patients manage their symptoms effectively at home, and providing guidance and decision support for primary care. We aim to establish a ‘gold standard’ of care by systematically analysing symptom clusters and current practices, iteratively improving pathways and systems of care, and working to disseminate better practices.

**Methods and analysis:** This mixed-method, multi-site study is informed by the principles of applied health services research, quality improvement, co-design, and learning health systems. It was developed in close partnership with patients (whose stated priorities are prompt clinical assessment; evidence-based advice and treatment; and help with returning to work and other roles) and with front-line clinicians. Workstreams and tasks to optimise assessment, treatment and monitoring are based in three contrasting settings: [1] specialist management in 10 long COVID clinics across the UK, via a quality improvement collaborative, experience-based co-design and targeted efforts to reduce inequalities of access; [2] patient self-management at home, with technology-supported monitoring; and [3] generalist management in primary care, harnessing electronic record data to study population phenotypes and develop evidence-based decision support, referral pathways and prioritisation criteria across the primary-secondary care interface, along with analysis of costs. Study governance includes an active patient advisory group.

**Ethics and dissemination:** LOCOMOTION is sponsored by the University of Leeds and approved by Yorkshire & The Humber - Bradford Leeds Research Ethics Committee (ref: 21/YH/0276). Dissemination plans include academic and lay publications, and partnerships with national and regional policymakers to influence service specifications and targeted funding streams.

**Study registration:** ClinicalTrials.gov: NCT05057260; ISRCTN15022307.

## Introduction

### What is long COVID?

We use the patient-made term ‘long COVID’^1^ to embrace the official categories of ‘ongoing symptomatic COVID-19’ (symptoms between 4 and 12 weeks) and ‘post COVID-19 syndrome’ (symptoms beyond 12 weeks) in any patient, irrespective of whether they were hospitalised or had a positive or negative SARS-CoV-2 test.^2^ Long COVID’s varied symptoms include fatigue, breathlessness, palpitations, dizziness, pain, neurocognitive dysfunction (‘brain fog’), sleep problems, exercise intolerance, functional disability in daily activities and reduced quality of life.^3-9^

Long COVID may be more common in people who had more severe acute illness or those with pre-existing conditions.^10 11^ Dysregulated immune response, immunothrombosis, endothelial dysfunction, multiple organ damage and dysautonomia all appear to play a role in its aetiology.^10 12 13^ Structural inequalities are important in the development and course of COVID-19 and may play a role in long COVID.^5 14-20^

People with lived experience of long COVID have a strong track record of contributing to the knowledge base on their condition. Long COVID was initially characterised by patients who came together in online communities after healthcare professionals (who thought of COVID-19 as a short-lived acute illness) had disbelieved or dismissed their stories.^1 21^ Many of those affected were health professionals themselves.^22^ Published patient-led research on long COVID includes large-scale symptom surveys,^18 23^ personal testimony and autoethnography, ^24 25^ co-design of services,^22 26^ and a manifesto for assessment and treatment.

### Long COVID management and services

Long COVID symptoms are characterised by symptoms and functional impairment that are multi-dimensional, episodic, and unpredictable in nature.^28^ The cornerstone of management is prompt multidisciplinary assessment and treatment with an emphasis on excluding serious complications, managing specific symptom clusters, and supporting whole-person rehabilitation.22 26 29 30 But it is unclear which patients need to be referred to specialist clinics and which tests and treatments are needed for whom.^11 31^ Recording of long COVID in primary care systems appears to be low.

As of March 2022, 1.5 million people in UK reported symptoms of long COVID.^33^ In 2021, NHS England invested £24 million to set up over 80 multidisciplinary long COVID clinics in England.^34^ Waiting lists for these specialist clinics are long and there are no equivalent services in Scotland, Wales or Northern Ireland. Specialist long COVID clinics vary in medical staffing levels, referral pathways, investigations and treatments. Demand exceeds supply, and the current demographics of clinic populations raise questions about inequalities of access in minority ethnic and other disadvantaged groups.

Whilst many long COVID patients could probably benefit from self-management resources (e.g. ‘Your COVID recovery’ website^35^), these are not currently well signposted or universally accessible, nor is there adequate guidance on how to support self-monitoring patients or when they may need escalation of care.

A new service specification expects primary care services to take on substantial elements of long COVID management.^36^ General practitioners—who claim chronic underfunding, understaffing, and task-shifting from secondary care—feel unable to cope with a new condition affecting large numbers of patients, especially in the absence of clear guidance and referral pathways.^19 37^

Given the numbers of people affected, the high levels of unmet need and burden of long-term disability, the disproportionate effect on disadvantaged people, the implications for the economy of long-term sickness absence, and the limited capacity in primary care, long COVID is an impending ‘system crisis’.^38^ There is an urgent need for applied research to optimise services and care pathways in a way that takes account of resource constraints and recognises the potential of patients themselves and generalist services to contribute to care, given appropriate support and resources.

## Methods and analysis

### Research aim

To optimise all aspects of long COVID care in UK, including access to services, care pathways and practices, and equity.

### Strategic objectives

Working in partnership with those who have lived experience of long COVID:

1. Establish a multi-site learning network of long COVID clinics and services across UK to capture and disseminate evidence-based practice.
2. Iteratively improve practices and protocols in long COVID clinics using a quality improvement collaborative and experience-based co-design to define best practice that ensures equitable access and care pathways.
3. Study self-management and symptom fluctuation in patients at home using a validated patient-reported outcome measure.
4. Use general practice electronic records to study population phenotypes and thereby develop evidence-based clinical templates, decision support, referral pathways and prioritisation criteria.
5. Evaluate the cost-effectiveness of different pathways.

### Research questions

Our research questions are shown in Box 1.

#### Box 1

Research questions

1. For specialist long COVID clinics:
  a. How can we use the quality improvement cycle to optimise the multidisciplinary assessment and care of people with long COVID?
  b. How can we draw on patients’ lived experience of services and the experience of front-line staff to inform this quality improvement?
  c. How can we improve access and reduce inequalities for disadvantaged and underserved groups?
  d. How can we best integrate clinical rehabilitation with vocational rehabilitation in the workplace?
  e. How can we optimise peer support for long COVID patients?
2. For patients at home:
  a. How can we monitor fluctuations and triggers in long COVID and use these data to aid self-management?
  b. How can we use patient-reported outcome measures to estimate condition severity, functional impact and quality of life in long COVID?
3. For integrated long COVID services across primary, secondary and community care:
  a. What referral and triage criteria are appropriate for referring patients to specialist services?
  b. What clinic investigations are appropriate for patients with specific symptoms or symptom clusters (e.g., chest pain, resting or exertional hypoxaemia, dysautonomia, symptoms suggestive of mast cell disorder, cognitive difficulties)?
  c. What is the effectiveness of specific interventions for patients with these and other relevant symptoms and symptom clusters?
  d. What are the appropriate skill mix and staffing levels for long COVID services of different types?
  e. What is the cost-effectiveness and healthcare utilisation of different care models?

### Study design

This mixed-method, multi-site study is co-designed with long COVID patients (whose stated priorities are prompt clinical assessment; evidence-based advice and treatment; and help with returning to work and other roles^1 22^) and front-line clinicians.

The study has three workstreams, each designed to optimise assessment, treatment and monitoring in a different setting (Figure 1). Workstream 1 addresses specialist management in 10 long COVID clinics. Workstream 2 addresses patient monitoring at home using a validated outcome measure (box 1). Workstream 3 addresses generalist management in primary care and pathways (including cost-effectiveness) across the primary/secondary care interface. A cross-cutting theme of patient and public involvement informs and supports all workstreams.

**Figure 1.**
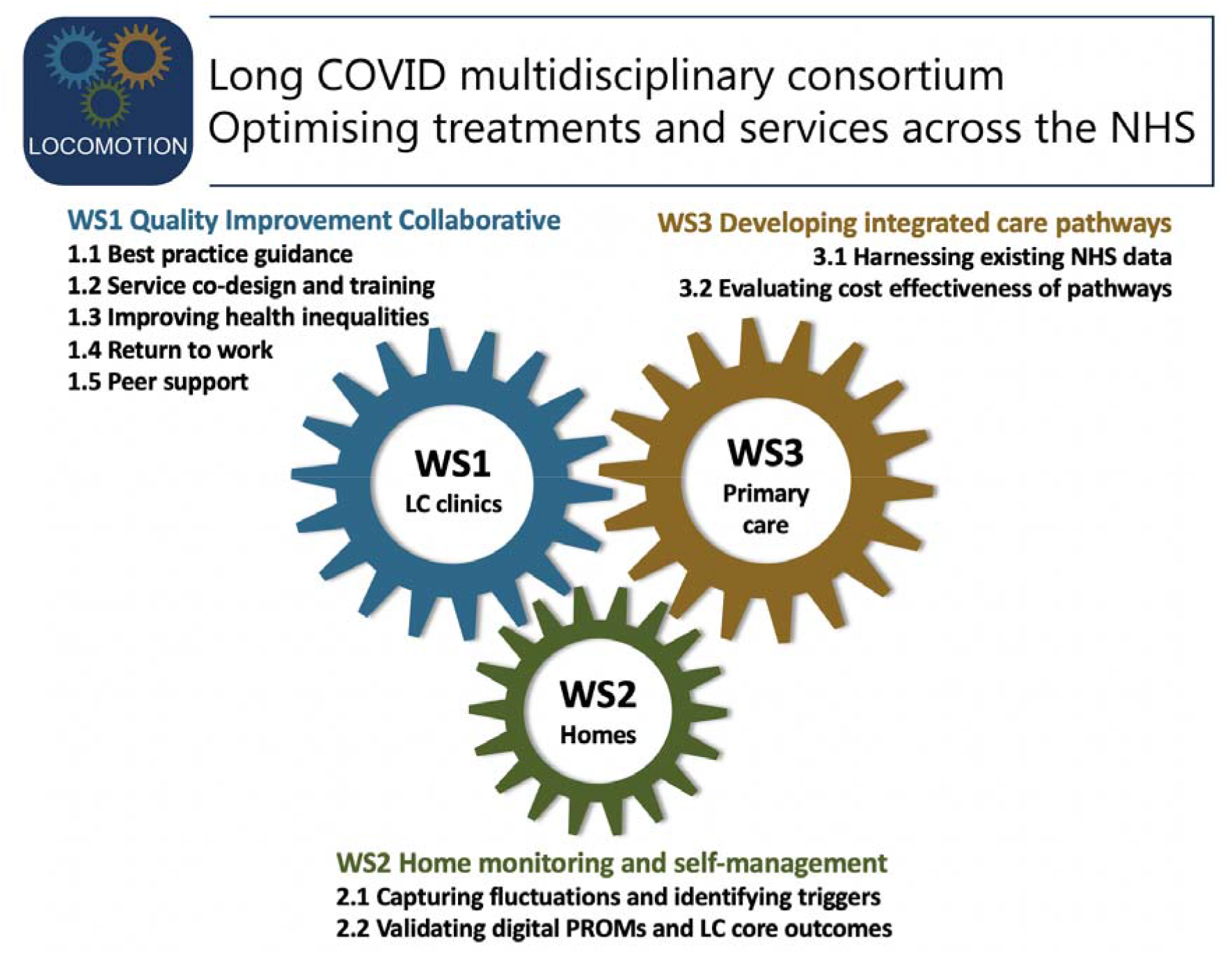
LOCOMOTION project

## Workstream 1: Long COVID specialist clinics

### 1.1 Multi-site quality improvement collaborative

Quality improvement collaboratives are networks of healthcare organisations that engage collectively in a cycle of data gathering, goal setting, action and evaluation, meeting regularly to compare findings and share resources.^39^ Although this model has had mixed success,^40^ if core principles are followed (e.g., good facilitation, clear goal setting, ensuring representatives from each organisation have clear channels for feeding their learning into acoordinated and strategic change effort),^39 41^ results can be dramatic.^41 42^

We will establish a quality improvement collaborative across 10 geographically and organisationally diverse long COVID services, with participation from site principal investigators, embedded clinician-researchers and patient partners. The collaborative will be chaired by an experienced clinician-researcher (TG) and will meet approximately monthly in a 2-hour video conference. Participants will share experience-based knowledge and research evidence, deliberate on best practice, and plan and evaluate practice change. The quality improvement cycle—prioritise a topic, set goals, identify data sources, implement change, collect data on performance and outcomes, then repeat—will be followed for both clinical (e.g. investigations, treatments) and more operational (e.g. referral criteria, service model, workforce) aspects of long COVID management.

### 1.2 Experience-based co-design

Experience-based [co-]design is an established improvement approach intended to ensure that health services are designed, redesigned, and improved around the needs and experiences of patients and front-line staff.^43^ Key features include a grounding in the perceptions and reactions of individual patients and staff, a focus on ‘emotional touch-points’ (points in the patient pathway that generate strong emotions such as frustration, anger, fear, or hopelessness). Experience-based co-design has a solid theoretical grounding in phenomenology (which, in this context, approximates to lived experience).^44^ This approach has been extensively applied in health service research and quality improvement.^45 46^

As a first step in the experience-based co-design method, a maximum-variety sample of approximately 15 adult patients and staff from each site will be interviewed about their experience of services using semi-structured or narrative interviews. Patient interviews will focus on emotional touch-points as these are likely to identify aspects of the service that need improvement.^47^ Staff interviews will capture ideas for service improvement. Interviews will be analysed thematically and fed back into local quality improvement.

### 1.3 Addressing inequalities

A qualitative sub-study will seek to understand and address the multiple intersecting inequalities in long COVID service utilisation.^12^ Working with and through participating NHS organisations, as well as community and advocacy organisations and selected social media outlets, we will recruit a maximum-variety sample of 30 people with long COVID representing key characteristics of underserved groups who have not [yet] been seen in a long COVID clinic. Sampling criteria include: poverty, homelessness, non-White ethnic groups (including Traveller communities), and those with disabilities; within all these groups we will seek to include a gender balance and recruit diverse age groups. In addition, we will interview a diverse sample of 15 key informants (individuals with relevant expertise on long COVID and health inequalities) via clinical, academic, policy and advocacy organisations. Interviews will explore symptom recognition (both self-reported and by clinicians), health-seeking behaviour, care pathways, motivations and disincentives to accessing healthcare support, and attitudes towards long COVID and stigma (e.g. relating to psychological symptoms). We will also explore emotional touch points, patient support networks and trajectories of care for those not receiving specialist long COVID healthcare.

All interviews will be transcribed, entered into a qualitative software package, and thematically analysed before being synthesised and fed into the work of the quality improvement collaborative (1.1 above), co-design (1.2 above), rehabilitation (1.4 below), home management (workstream 2), and trajectories of care and pathway redesign (workstream 3).

### 1.4 Vocational rehabilitation

People living with long COVID can find it difficult to return to work, and those who have been able to return to work are experiencing work instability (defined as is a mismatch between an individual’s abilities and the demands of their job^48^). We will explore the needs of 20 working-age people recruited from long COVID clinics and selected for diversity in age, gender, ethnicity, occupation and career stage to inform the development of appropriate return-to-work programmes (vocational rehabilitation pathway). To understand return-to-work policies and procedures, we will interview a purposive sample of occupational health, human resource and managerial professionals from a representative range of organisations, recruited via social media and researchers’ local contacts. Based on these interviews and our experience of designing vocational rehabilitation programmes for other acute-onset, long-term conditions, we will develop an individually tailorable, co-ordinated programme of support, education and advice. This advice will be relevant for people with long COVID, their family and others involved in the person’s vocational role such as employers and disability employment advisors. We will test the proposed programme for acceptability and feasibility with a further sample of 20 diverse participants, as well as clinicians and therapists.

### 1.5 Peer support

Long COVID was first characterised in online peer support groups and several participating sites have local peer support groups but it is not known how best to structure and support such groups. We will undertake a hermeneutic literature review of peer support models in comparable conditions (e.g. chronic pain) and conduct interviews with stakeholders involved in delivering or supporting peer support for long COVID. We will share findings with workstream 1.1. Through discussion, we will identify which features of successful peer support are relevant to long COVID and consider how to improve existing models (if present) or establish new peer support services. Using the quality improvement cycle, we will collect data to evaluate and improve as the peer support groups evolve.

## Workstream 2: Home monitoring and self-management by long COVID patients

### 2.1 Monitoring fluctuations, symptoms and associated triggers

Long COVID is characterised by fluctuating and difficult to manage symptoms with high variability between individuals. Its relapsing and remitting presentation may be exacerbated by triggers, though patients report difficulty with pacing strategies, and little is known about what causes fluctuations or the nature of triggers. We propose disordered relationships in symptoms and activities are underpinned by possible pathologies across multiple body systems including the central nervous system and interoceptive pathways.

This sub-study uses an intensive longitudinal design. Some of the key COVID-19 Yorkshire Rehabilitation Scale questions (Box 2) have been embedded into a bespoke digital platform which monitors general health, symptom fluctuations, and potential triggers, and is linked to wearable sensors. We hypothesise that physical, cognitive and emotional triggers will predict symptoms experienced at subsequent timepoints the same day or day after. The relationship between triggers and symptoms will vary within and between individuals and bespoke understanding is required for effective self-management.

#### BOX 2

The COVID-19 Yorkshire Rehabilitation Scale: A validated patient-reported outcome measure for long COVID

We have previously developed the Covid-19 Yorkshire Rehabilitation Scale, the first patient-reported outcome measure for long COVID. This instrument is recommended in the National Institute for Health and Care Excellence (NICE) rapid guideline for long COVID and NHS England service guidance and adapted in the WHO self-management booklet. ^2 49-51^ The scale has been digitised by a private digital health company (ELAROS). The patient completes the questionnaire on a smartphone application and the clinicians access the results on a web portal and both use the system to monitor progress, fluctuations and response to ongoing treatments for long COVID.

The original COVID-19 Yorkshire Rehabilitation Scale is a 23-item patient-reported outcome measure which grades the severity of key symptoms, functional limitations, overall health and additional symptoms on an 11-point Likert scale and also captures pre-COVID scores for comparison. Questions 1-10 form the symptom severity subscale (score 0-100), 11-15 the functional disability subscale (0-50), 16 is the overall health score (0-10) and 17-23 the additional symptoms subscale (0-60). The psychometric analysis in a sample of 187 long COVID patients showed good data quality, satisfactory scaling and targeting and good reliability both overall (Cronbach’s alpha 0.891) and for individual subscales.^53^

Initial testing of a previous version of the COVID-19 Yorkshire Rehabilitation Scale in 370 community patients from a single long COVID clinic (Leeds) appears to reveal three clinical severity phenotypes (mild, moderate and severe) for both individual symptom clusters and functional disability (Figure 1)^12^. Such condition severity phenotypes with remitting relapsing nature of the condition suggest common mechanisms driving the array of symptoms but this is yet to be fully established.

Preliminary Rasch analysis of the original version of COVID-19 Yorkshire Rehabilitation Scale revealed dysfunctional scale response categories. The instrument was modified by replacing the 11-point Likert scale with a simpler 4-point scale. This version will be made available on the digital platform for the LOCOMOTION study so it can be used to gather individual data for clinical assessment and also present these data in pseudonymised form for aggregated analysis (e.g. to derive psychometric properties of the scale).

A full version of the modified COVID-19 Yorkshire Rehabilitation Scale is available in the appendix on bmj.com [see supplementary file 1].

We will recruit a diverse sample of 400 patients who are awaiting their initial long COVID appointment. We will conduct a time-series study (referral, 6 weeks, 12 weeks) comprising brief symptom surveys six times daily for a 7-day period, along with sensor and accelerometer data plus self-reported activities and emotional triggers. This daily ecological momentary assessment includes continuous data collection of the following using activity sensors (Axivity): physical activity levels (step count, intensity of physical activity, timings and duration of physical activity), sleep (timing and duration). Participants will also answer a series of questions throughout the day about their sleep, daily activities, symptoms and symptom impact, post-exertional malaise, stress and anxiety.

We will also recruit 50 participants from one site (Oxford) who will additionally be monitored to explore the potential for using heart rate and heart rate variability in biofeedback therapy. Physiological phenotyping will include the COVID-19 Yorkshire Rehabilitation Scale plus additional measures of heart rate and heart rate variability (using a wrist or chest-worn sensor—Fitbit Sense and Polar H10).

Part-way through the main workstream 2 study, a small qualitative evaluation will be undertaken using semi-structured interviews with 20 patient and 10 clinician participants. For patients this will include reflections on their COVID experience, a think-aloud exercise informed by theories of technology usability (dimensions of efficiency, effectiveness and satisfaction), and suggestions for improving the app or the study. Data will be analysed thematically using insights from sociotechnical theories.

Findings from 2.1 will feed into the quality improvement collaborative (workstream 1.1 above) and the pathway redesign (workstream 3.1 below).

### 2.2 Use the COVID-19 Yorkshire Rehabilitation Scale to monitor symptoms in clinic cohorts

Preliminary validation data from the COVID-19 Yorkshire Rehabilitation Scale, a condition-specific outcome measure for Long COVID,^54^ suggest that patients scoring low on one symptom tend to score low on other symptoms too (see Figure 2)^12^ — consistent with severity phenotypes seen in the post-hospitalisation COVID-19 study (PHOSP-COVID),^10^ and suggesting that long COVID may be driven by common mechanisms affecting multiple body systems. Workstream 2.2 will test this hypothesis to determine the extent to which the COVID-19 Yorkshire Rehabilitation Scale can be used for triage, targeting interventions and capturing the response to treatments.

**Figure 2.**
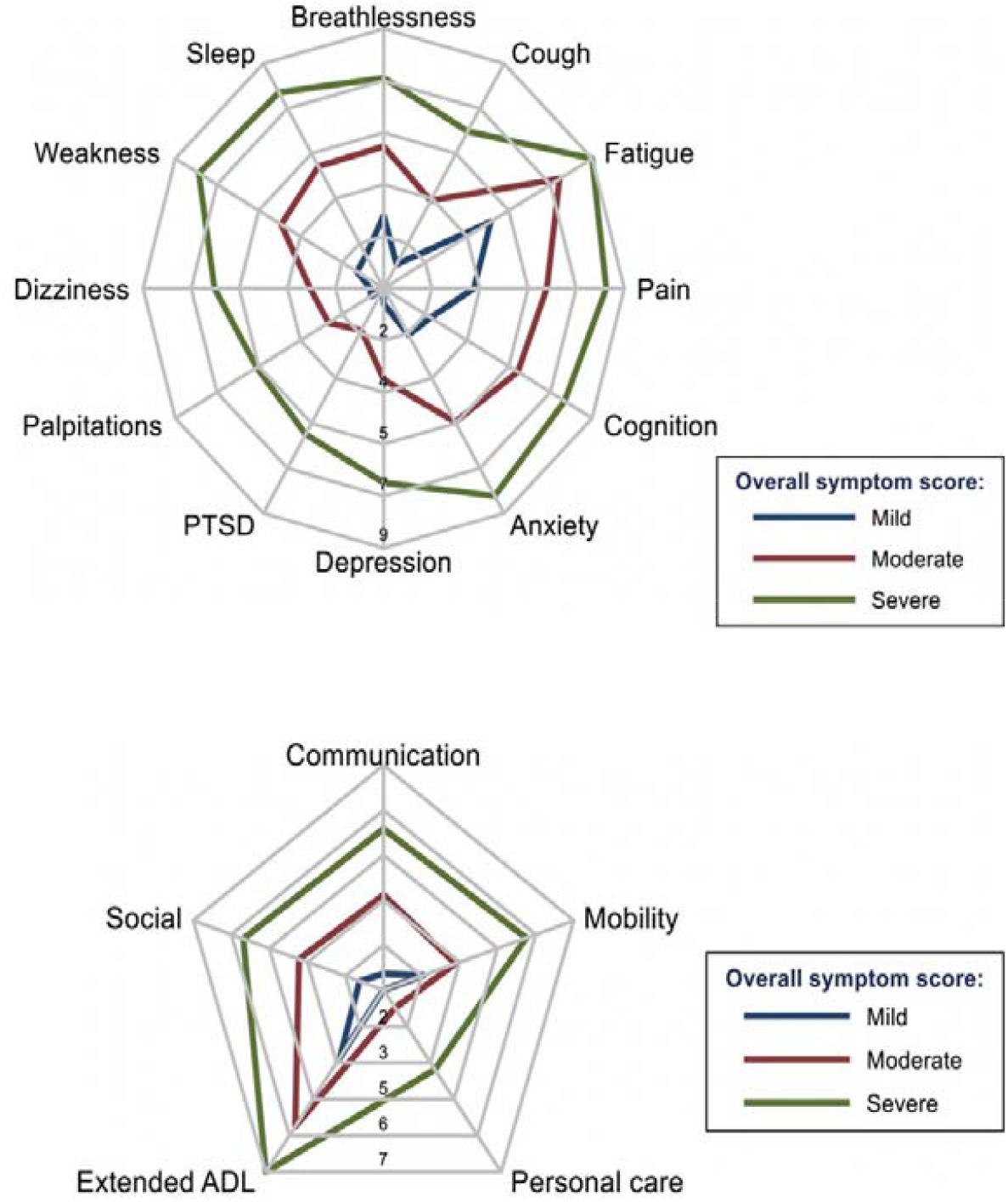
Aggregate scores from a clinic population on the COVID-19 Yorkshire Rehabilitation Scale patient reported outcome measure a) Mean symptom severity score of 370 patients, plotted as three subgroups (severe >6, moderate 3-5.9 and mild <3). Since radar plots do not intersect, these preliminary data suggest a single syndrome rather than several different syndromes of long COVID b) Mean functional ability scores on same sample Figure reproduced with permission from Sivan et al.^12^

Longitudinal COVID-19 Yorkshire Rehabilitation Scale data on a large sample of long COVID clinic patients will be collected quarterly from participants using the ELAROS digital app and web portal^52^ or equivalent. The COVID-19 Yorkshire Rehabilitation Scale has been modified based on increasing knowledge of the condition and preliminary psychometric analysis.^55^ Further psychometric evaluation of the modified COVID-19 Yorkshire Rehabilitation Scale in this new sample will use a Rasch measurement model to explore the scale item’s model fit, local dependency, response category functioning and differential item functioning. If necessary, we will refine the scale through an iterative process of psychometric testing and modifications. Each Rasch scale assessment will use data from between 500 and 600 patients who have completed the COVID-19 Yorkshire Rehabilitation Scale as part of their standard assessment procedures across our ten clinics. We anticipate iterations to the scale during the project depending on findings of the Rasch scale assessment. When a stable item set is decided, the total available sample will be used to provide final scale calibrations.

The digital platform will also capture other aspects of long COVID using symptom-specific patient-reported outcome measures and quality of life measures. Data from patient completed COVID-19 Yorkshire Rehabilitation Scale and EuroQoL EQ-5D questionnaires will be collected from all sites uniformly and each site can capture other measures in the digital platform if they wish. The platform will also have the ability to include the World Health Organisation’s core set of outcome measures that is currently being developed.^56^ This work will enable testing of a core set of measures that can capture this new condition comprehensively and compare with the WHO’s International Classification of Functioning Disability and Health (ICF) framework.^57^

## Workstream 3: Developing and evaluating long COVID integrated care pathways

### 3.1 Pathway development

We will use both retrospective and prospective cohorts to answer research questions 3a-3d in Box 1. For patients newly referred to long COVID clinics, we will collect prospective data (to inform national standards for routine data collection moving forward) and link to clinical information captured in routine care. In a second cohort of patients suspected of having long COVID but not referred to long COVID clinics, routine data capture from healthcare provision will be used to understand pathways and demographic factors to increase appropriate referrals moving forward. An integrated dataset will be constructed from primary care, community care and specialist long COVID clinics to develop and evaluate effective and cost-effective service models which incorporate the quality principles and best practice guidance developed in workstream 1.1 above.

We will use advanced analytical techniques (such as latent class analysis and clustering) pertinent to providing insight into long COVID phenotypes based on pseudonymised patient-level variables, including patient outcomes and co-morbidities. These patient groupings will then be analysed in the context of healthcare settings, using network analysis to understand the effect of patient trajectories, enabling validation or modification of existing guidelines, leading to implementation of new clinical decision support tools in electronic record systems.

We will use approved mechanisms for data linkage via NHS Digital and work within established trusted research environments. Data linkage for practice data and patient clinic data for Imperial will take place in North West London Whole Systems Integrated Care, Salford’s Integrated Record for Salford, and via the Oxford-Royal College of General Practitioners Clinical Informatics Digital Hub (ORCHID) for all other sites.^58^ Brief details of these datasets are shown in Box 3 and further detail given in the appendix on bmj.com. All research analysis will be undertaken on de-identified data.

#### Box 3

Data sets that will contribute towards modelling long COVID phenotypes and cost-effectiveness of treatments

Data resource 1: iCARE/North West London Whole Systems Integrated Care is a Heath Data Research UK Alliance Trusted Research Environment and Salford - unique health management platforms covering a population of 2.6 million in NW London and the Salford Integrated Record (includes the ICHT and Salford long COVID clinics).

Data resource 2: The Oxford-Royal College of General Practitioners Research and Surveillance Centre is one of Europe’s oldest sentinel networks and recruited to be nationally representative. It now consists of a network of in excess of 1800 practices’ data (N>18million, 32% of the English national population). Data from this Research and Surveillance Centre, as well as other data from across LOCOMOTION are being made available via ORCHID (http://orchid.phc.ox.ac.uk), a Health Data Research UK listed trusted research environment and meets NHS Digital’s Data Security and Protection requirements. Researchers across LOCOMOTION with an approved analysis requirement can apply to access data remotely within ORCHID. Data goes through a privacy protecting statistical disclosure control process prior to leaving the ORCHID trusted research environment. ORCHID is supporting national COVID-19 surveillance of the UK Health Security Agency and four national core studies.^60^ ORCHID is a pseudonymised dataset using an NHS Digital approved method allowing data to be linked to national test results, immunisation, emergency care, hospital, and death datasets at an individual patient level.

Data resource 3: National General Practice Data for Research and Planning via the National Core Studies Portal.

### 3.2 Pathway cost-effectiveness analysis and model testing

This workstream aims to assess the cost-effectiveness of different care pathways and develop and test efficient service models. Service models vary substantially across England.^61^ Comparing these using standardised health economic outcome measures will enable an understanding of the cost-effectiveness aspects of service delivery.

Following established frameworks for model conceptualisation we will incorporate collaborative stakeholder and public involvement,^62 63^ and address known methodological challenges for public health economic modelling.^64^ We will evaluate the cost-effectiveness of alternative care pathways from a societal perspective, incorporating health and social care costs incurred by the public sector as well as productivity losses and out-of-pocket expenditures. The primary analysis will assess the cost-effectiveness of alternative strategies in terms of incremental cost per quality-adjusted life year gained. We will use cost-effectiveness thresholds of £20-30,000 and £60,000 per quality-adjusted life year gained.^65 66^ Secondary analysis will evaluate cost-consequences of alternative care pathways and disaggregate these in terms of disparate outcomes such as the proportions of individuals who return to pre-COVID levels of productivity and those who continue to experience excessive out-of-pocket expenditures.^67^

We will validate models using real-world data from existing platforms (Data resources 1-3, box 3).^68^ Probabilistic sensitivity analysis will assess the impact of uncertainties of all incorporated parameters.^69^ We will use cost-effectiveness acceptability curves to show the probability of cost-effectiveness of each evaluated strategy at alternative cost-effectiveness thresholds held by decision-makers. Scenario analyses will assess the impacts of different model structural assumptions.

## Equality, diversity and inclusion

Meeting the needs of all patients with long COVID is at the heart of the LOCOMOTION study. Specifically, workstream 1.3 is dedicated to exploring the experiences of underserved population groups. This workstream will identify key facilitators and barriers to delivering good quality healthcare to these communities and findings will feed into co-design of long COVID clinics, understanding the features of Long COVID and validating Patient Reported Outcome Measures. In addition, workstreams exploring vocational rehabilitation, home management, peer support and care pathway redesign will specifically gather and analyse data relating to populations most at risk of long COVID.

To further understand the disproportionate impact of long COVID across the population, we created a seven-member over-arching patient advisory group inclusive of diverse cultural, ethnic and socioeconomic groups. This group contributes to the governance of LOCOMOTION and provides overall management of patient and public involvement, which is embedded throughout the study. Members of the group link into each workstream to ensure relevant and meaningful involvement activity within each one. The advisory group has worked with all workstreams to maximise inclusivity and reduce exclusion by design. Revisions to plans include widening accessibility to the COVID-19 Yorkshire Rehabilitation Scale and EuroQol EQ-5D patient reported outcome measures and supporting alternative formats for interviews where digital/remote attendance may be difficult.

To support recruitment across all workstreams we will offer translation services on an individual basis where required to facilitate qualitative interviews and the consent process. This will include translation of patient information sheets, consent forms and outcome measures and use of bilingual researchers or interpreters as necessary. For participants not currently receiving specialist support for long COVID, we will also try to offer incentive payments in recognition of the barriers to engaging with research experienced by underserved groups.^70^

## Patient and public involvement

For the outputs of LOCOMOTION to meet patient need the study must be responsive to the patient voice that lies at the centre of its design, development and delivery. Members of the patient advisory group attended proposal research planning meetings and met separately to analyse and develop the research aims, objectives, and questions, ensuring these align with the key research priorities of those with long COVID. All advisory group members have lived experience of long COVID. Some also have experience of design and evaluation of research bids and policy task forces. They have contacts with wider patient community groups. The advisory group meets quarterly to review progress, ensure the research continues to answer relevant issues and that findings can inform long COVID care. In addition, two patients from each of the ten long COVID NHS clinics will form the patient advisory network, which will liaise with the advisory group, providing local intelligence about long COVID experiences and clinic access. The advisory group and advisory network are supported by an experienced Patient and Public Involvement/Engagement Manager. The patient-level and service-level measures have been determined by patients, healthcare professionals and researchers using consensus methods (Table 1 and 2).

**Table 1:**
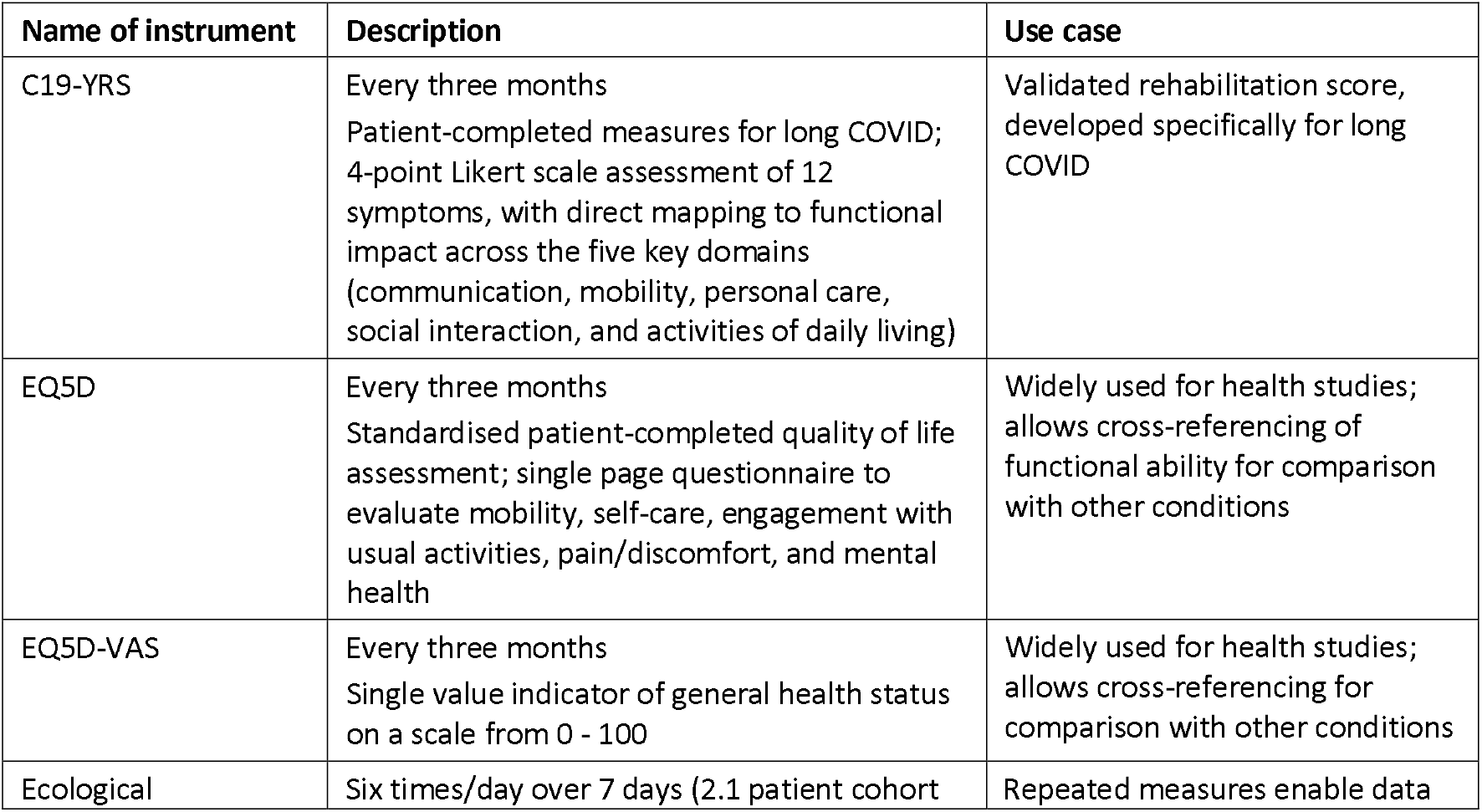
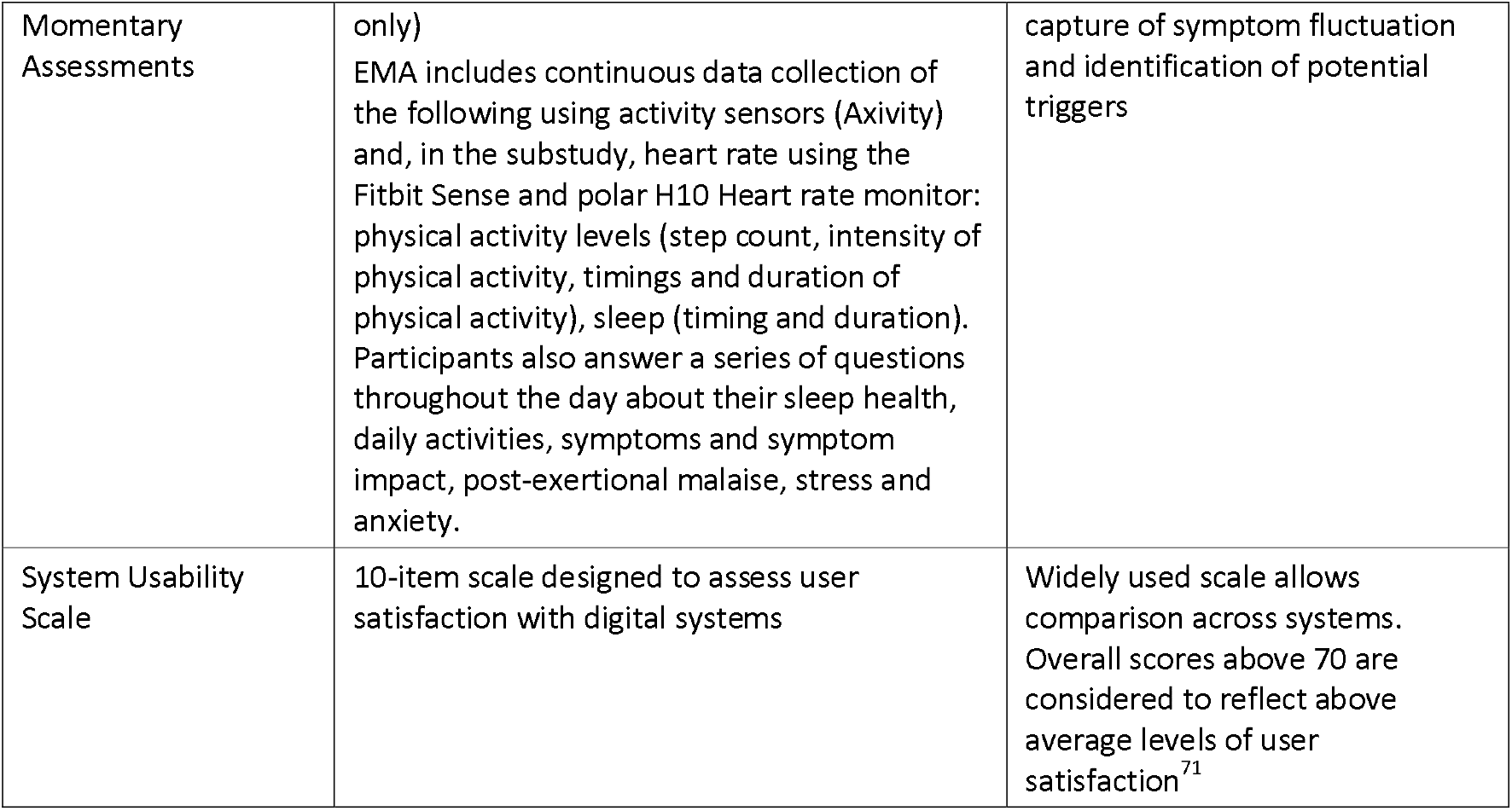
Patient-level measures to be used in LOCOMOTION sites

**Table 2:** Service-level measures to be used in LOCOMOTION sites

| Measure | How measured | Type of data |
| --- | --- | --- |
| Evidence of patient-focused iterative change to clinic services | Ongoing data collection throughout the study from interactions with sites (PIs, research fellows, patients, and local multi-disciplinary teams) and simple summary statistics will provide evidence of changes to and evaluation of long COVID clinic services | Qualitative and Quantitative |
| Patient experiences of efforts to reduce inequalities | Interviews with patients and key informants will explore symptom recognition, health-seeking behaviour, care pathways, motivations/disincentives to accessing healthcare support, attitudes towards long COVID and stigma. | Qualitative |
| Patient experiences of tailored vocational rehabilitation as per guidelines | Qualitative data from interviews with patients, professionals involved in long COVID clinics and key informants will explore the impact of long COVID on return-to-work and job retention, including access to and from work and within work, adaptations required for work. | Qualitative |
| Cost per quality-adjusted life year QALY | Cost-effectiveness of alternative models of service delivery will be expressed in terms of incremental cost per QALY | Quantitative |
| Cost-effectiveness acceptability curves | Cost-effective acceptability curves will be used to show the probability of cost-effectiveness of each of the evaluated strategies at alternative cost-effectiveness thresholds held by decision makers | Quantitative |

## Statistical analysis

The rationale for sample size for the tasks involving quantitative data (2.1, 2.2, 3.1, 3.2) is uploaded as a separate document [see supplementary file 3]

## Ethics and dissemination

Ethics approval was obtained from Bradford and Leeds Research Ethics Committee on behalf of Health Research Authority (HRA) and Health and Care Research Wales (HCRW) on 06 Jan 2022 (reference: 21/YH/0276). Retrospective data required for General Practice and Hospital Episode Statistics linkage cannot be provided in anonymised form. These data will be collected under the Control of Patient Information (COPI) notice initially and subsequently under authorisation from the Health Research Authority Confidentiality Advisory Group.

Dissemination will include both academic publications and lay summaries in various formats. Relevant long COVID phenotypes will be published in the Health Data Research UK National Phenotype Library, together with their computable definition, and assigned a DOI. Policy impact will be aided by our strong existing links to NHS England, and by the fact that several of the co-investigators are on the UK Long COVID National Task Force. Dr Sivan, who co-leads LOCOMOTION, is advisor for the World Health Organisation (Europe) on COVID-19 rehabilitation and is also involved in the World Health Organisation working party to develop a core set of outcome measures for long COVID.

We have links with other long COVID projects based in the UK and beyond to enable co-learning and maximise impact. In particular, our links with the Symptoms, Trajectory, Inequalities and Management: Understanding Long-COVID to Address and Transform Existing Integrate Care Pathways (STIMULATE ICP) study (https://www.stimulate-icp.org/) will enable sharing and evaluating clinic data for exploring mechanisms and developing treatment algorithms. Links with the Therapies for Long Covid platform^72^ will facilitate development of condition specific measures for long COVID and compare the psychometric properties of these new measures.

## Conclusion

Long COVID is an emerging health condition with significant associated morbidity for large numbers of people and disproportionate impact on particular social groups. Two years since global medical communities were first faced with patients with acute COVID-19 infection, we have a number of effective treatments and a suite of vaccines. However, our understanding of long COVID remains poor, community health services are severely stretched, and are struggling with the growing burden of chronic illness following COVID-19 infection. There is an urgent need to develop evidence-based treatments that address multimorbidity and inequalities. Long COVID is an example of how integrated service delivery approaches should be designed.

Addressing the need for rapid learning from real-world multidisciplinary care,^4^ the LOCOMOTION study will combine data modelling and home monitoring with analysis of lived experiences to recognise and support effective management of long COVID. We will focus on capturing patients’ multisystem symptoms and rehabilitation needs and providing individualised care programmes that aim for medical management and a functional improvement, including (where appropriate) return to work.

## Data Availability

We will use Open Science Framework (OSF) to share of all research outputs, including
data, codes, and other types of information that has the potential to aid the advancement
of scientific progress and benefit other researchers by adding transparency to the
research process. Data items from individual studies will be shared in relevant consortium
institutional repositories (the University of Leeds public data repository for studies led by Leeds) to increase exposure. The OSF will consist of two levels: a data dictionary with basic info about the study, and the more detailed dataset (for further analysis / meta-analysis). Data will be issued with a Digital Object Identifier (DOI) which will allow it to be referenced and to make it easier for others to identify and access relevant files.

## Data statement

We will use Open Science Framework (OSF) to share of all research outputs, including data, codes, and other types of information that has the potential to aid the advancement of scientific progress and benefit other researchers by adding transparency to the research process. Data items from individual studies will be shared in relevant consortium institutional repositories (e.g., the University of Leeds’s public data repository for studies led by Leeds) to increase exposure. The OSF will consist of two levels: a data dictionary with basic info about the study, and the more detailed dataset (e.g., for further analysis / meta-analysis). Data will be issued with a Digital Object Identifier (DOI) which will allow it to be referenced and to make it easier for others to identify and access relevant files.

## Supplementary files

1 Modified Covid Yorkshire Rehabilitation Scale (C19-YRS) questionnaire

2 LOCOMOTION statistical analysis

