## Supplementary File 2 for "Protocol for mixed-method study by LOng COvid Multidisciplinary consortium: Optimising Treatments and servIces acrOss the NHS (LOCOMOTION)"

### Statistical appendix

Task 2.1:

For the main workstream we will recruit 400 participants with the anticipation of a 30% dropout, giving 300 patients (40 per site) using consecutive sampling. For the single site sub-study we will invite 50 participants, allowing for dropouts and leaving approximately 30 participants to ensure meeting central limit theorem criteria for reliability analysis and between 25-50 for meeting feasibility criteria.[1]

Task 2.2:

There are no fixed or established guidelines on the sample size required for the psychometric testing in this task. Each analysis will vary depending upon its specific purpose. A sample size of between five and six hundred is robust to obtain a confidence level of 99% for item calibrations to be stable within less than half a logit,[2] although additional parameters also need to be considered, such as targeting and item-bias testing group sizes.

Larger sample sizes (eg approximately 5000 across the ten clinics) will provide more stable calibrations, making them less susceptible to random variations within the data. Larger sample sizes also allow for a cross-validation methodology (splitting the sample and replicating the analysis), reducing the risk of incorporating random error into final conclusions.

Patients will be excluded from the analysis if they have missing data for more than 50% of the main-body YRS items to reduce large errors and fluctuations.

Tasks 3.1 and 3.2:

There is no basis for calculating a sample size at this point in time as analyses will be hypothesis-driven, iterative and exploratory, responding to WS1. With over 7 million patients across the datasets, we have sufficient data.
